# Systematical assessment of the impact of single spike mutations of SARS-CoV-2 Omicron sub-variants on the neutralization capacity of post-vaccination sera

**DOI:** 10.1101/2023.09.08.23295177

**Authors:** Maeva Katzmarzyk, Denise Christine Clesle, Joop van den Heuvel, Markus Hoffmann, Henk Garritsen, Stefan Pöhlmann, Henning Jacobsen, Luka Cicin-Sain

## Abstract

The evolution of novel SARS-CoV-2 variants significantly affects vaccine effectiveness. While these effects can only be studied retrospectively, neutralizing antibody titers are most used as correlates of protection. However, studies assessing neutralizing antibody titers often show heterogeneous data. To address this, we investigated assay variance and identified virus infection time and dose as factors affecting assay robustness. We next measured neutralization against Omicron sub-variants in cohorts with hybrid or vaccine induced immunity, identifying a gradient of immune escape potential. To evaluate the effect of individual mutations on this immune escape potential of Omicron variants, we systematically assessed the effect of each individual mutation specific to Omicron BA.1, BA.2, BA.2.12.1, and BA.4/5. We cloned a library of pseudo-viruses expressing spikes with single point mutations, and subjected it to pooled sera from vaccinated hosts, thereby identifying multiple mutations that independently affect neutralization potency. These data might help to predict antigenic features of novel viral variants carrying these mutations and support the development of broad monoclonal antibodies.

## Introduction

Since its emergence in late 2019, the severe acute respiratory syndrome coronavirus 2 (SARS-CoV-2) has rapidly spread globally, posing a great risk for public health. Global vaccination campaigns against the coronavirus disease 2019 (COVID-19) greatly increased anti-SARS-CoV-2 immunity in the general population, which simultaneously restricted viral spread and enhanced selection pressure on the virus. As a consequence of vaccination and/or infection-induced anti-SARS-CoV-2 immunity, SARS-CoV-2 has undergone antigenic drift and concomitantly, novel SARS-CoV-2 variants emerged that gained the ability to evade acquired immunity. The impact of mutations on vaccine performance against viral variants is best studied in clinical studies, but these are by their nature retrospective analyses and cannot provide timely information about immune escape risks of emerging variants. Therefore, serological correlates of vaccine efficiency, such as neutralizing antibody titers, have been used to prospectively assess the expected immune protection against novel variants of concern. In line with clinical evidence, vaccination- or infection-elicited antibodies efficiently neutralize initial, antigenically similar variants, but the antigenically distant Omicron variant and its increasing number of continuously evolving sub-variants escape antibody neutralization (Cao et al., 2023; Pather et al., 2023). However, published neutralization data are often heterogeneous and difficult to compare across studies, limiting the translational value of these data. Namely, technical differences in laboratory protocols and procedures are likely to affect the robustness of neutralization data and thereby limit the comparability of neutralization efficiencies across studies.

The gene of the spike (S) protein of the original Omicron BA.1 variant carries a high number of mutations (31 amino acid exchanges, 3 short deletions and one insertion of 3 amino acids), which may have affected not only the immune escape, but also increased its transmissibility while reducing disease severity (J. Chen et al., 2022; Peacock et al., 2022; Sievers et al., 2022; Wolter et al., 2022). Several of these mutations, especially amino acid substitutions in the spike protein, are shared with previous variants of concern, like D614G, N501Y, K417N, and E484K/A. The insertion of three amino acids to the spike protein, ins214EPE, however, was observed for the first time in the Omicron lineage, along with numerous other mutations (Venkatakrishnan et al., 2021). Such antigenic drifts significantly affected virus characteristics like enhanced escape from antibody neutralization, thereby increasing the risk of re-infection even across different Omicron sub-variants (Cameroni et al., 2022). The Omicron BA.2 (B.1.1.529.2) variant has acquired additional mutations mainly in the N-terminal domain (NTD) and the receptor binding domain (RBD) (Yu et al., 2022). Omicron BA.2 rapidly became the globally dominant variant just shortly after the emergence of Omicron BA.1 (Yu et al., 2022). BA.2.12.1, a sub-lineage of Omicron BA.2, emerged in the United States beginning of June 2022 with novel spike mutations L452Q in the RBD and S704L as a mutation of the fusion machinery (Bowen et al., 2022). One early Omicron sub-variant that is sparsely characterized compared to other variants is Omicron BA.3, which shows no novel spike mutations, but a combination of the mutations found in BA.1 and BA.2 (Desingu et al., 2022). While the above-mentioned Omicron sub-variants all show a comparable immune escape from index-specific antibodies with a 4.2-fold to 12.5-fold decrease in neutralization capacity relative to index in boosted subjects, the sub-variants BA.4 and BA.5 that emerged in summer 2022, showed another significant increase in antigenic distance with an average 22.6-fold decrease in serum neutralization by boosted subjects (Jacobsen et al., 2022).

Omicron BA.4 and BA.5 share an identical spike protein sequence, including two novel mutations (L452R and F486V) in the receptor-binding motif that were not seen in previous Omicron sub-variants. The BA.5 variant quickly spread globally and was the last dominating variant in Germany, from June to late 2022, indicating additional factors favoring viral fitness of BA.5 over BA.4 other than the mutational profile of the spike glycoprotein. In late 2022, new sub-variants such as BQ.1, BQ.1.1, or XBB.1.5 emerged, and even booster vaccinations with vaccines adapted to Omicron BA.5 or hybrid immunity by BA.5 breakthrough infection just slightly enhanced neutralization titers to these newly emerging Omicron sub-variants which contained only a few additional novel spike mutations (Chen et al., 2023; Planas et al., 2022; Qu et al., 2022; Yamasoba et al., 2023). In addition, single RBD amino-acid substitutions that were highly frequent in several SARS-CoV-2 infection waves are able to drive immune evasion as observed in serum from boosted and BA.2 or BA.5 breakthrough infected individuals (S. Chen et al., 2023). Therefore, few mutations in strategic positions may significantly increase neutralization escape, but the contribution of each individual mutation remains unclear. It is reasonable to assume that SARS-CoV-2 will continue to evolve, and that genetic drift will lead to new viral variants able to evade population immunity. A thorough understanding of mutations that independently affect immune escape might help to predict their effects on the magnitude of immune escape by emerging variants featuring them in novel combinations. Moreover, the design of monoclonal antibodies as well as the assessment of convalescent plasma pools for therapeutic use would benefit from a better understanding of these mutations.

In this study, we first optimized the robustness of pseudo-virus neutralization assays by defining virus input titer and incubation time after infection that result in the lowest technical variance. We then performed optimized neutralization assays with full spike pseudo-viruses of clinically relevant, as well as newly emerging Omicron sub-variants including BA.2.75, BA.4.6, BA.5.9, BF.7, BQ.1, BQ.1.1, and XBB.1.5, to get an initial understanding of the variant-specific potential to escape neutralizing antibodies. Finally, we generated a library of each Omicron-associated spike mutation from BA.1, BA.2, BA.2.12.1, BA.3, and BA.4/5 and systematically characterized their specific impact on serum neutralization. Omicron-associated single mutations were cloned into pseudo-viruses carrying the SARS-CoV-2 index spike protein and analyzed in the optimized pseudo-virus neutralization assay, using sera of monovalent vaccinated, and SARS-CoV-2 infection-naive individuals to define the effect of each single omicron-associated mutation on neutralization escape from polyclonal, vaccine-elicited immune responses.

## Methods

### Systematic literature research

The systematic literature research was performed in MEDLINE via PubMed including all published and peer-review studies up to December 31^st^ 2022. The following search term was used: “SARS-CoV-2 AND *mutation* AND neutralization assay”. For the term “mutation” each single Omicron-associated spike mutation was inserted. Studies meeting the following inclusion criteria were included: published studies with neutralization data on clinical specimen using SARS-CoV-2 viruses with single Omicron-associated spike mutations in the index spike background. Studies assessing single spike mutations that were cloned in the D614G spike background were included likewise. Studies that did not fit the inclusion criteria were excluded. An overview of the study identification and selection process is provided in figure 1.

**Figure 1:**
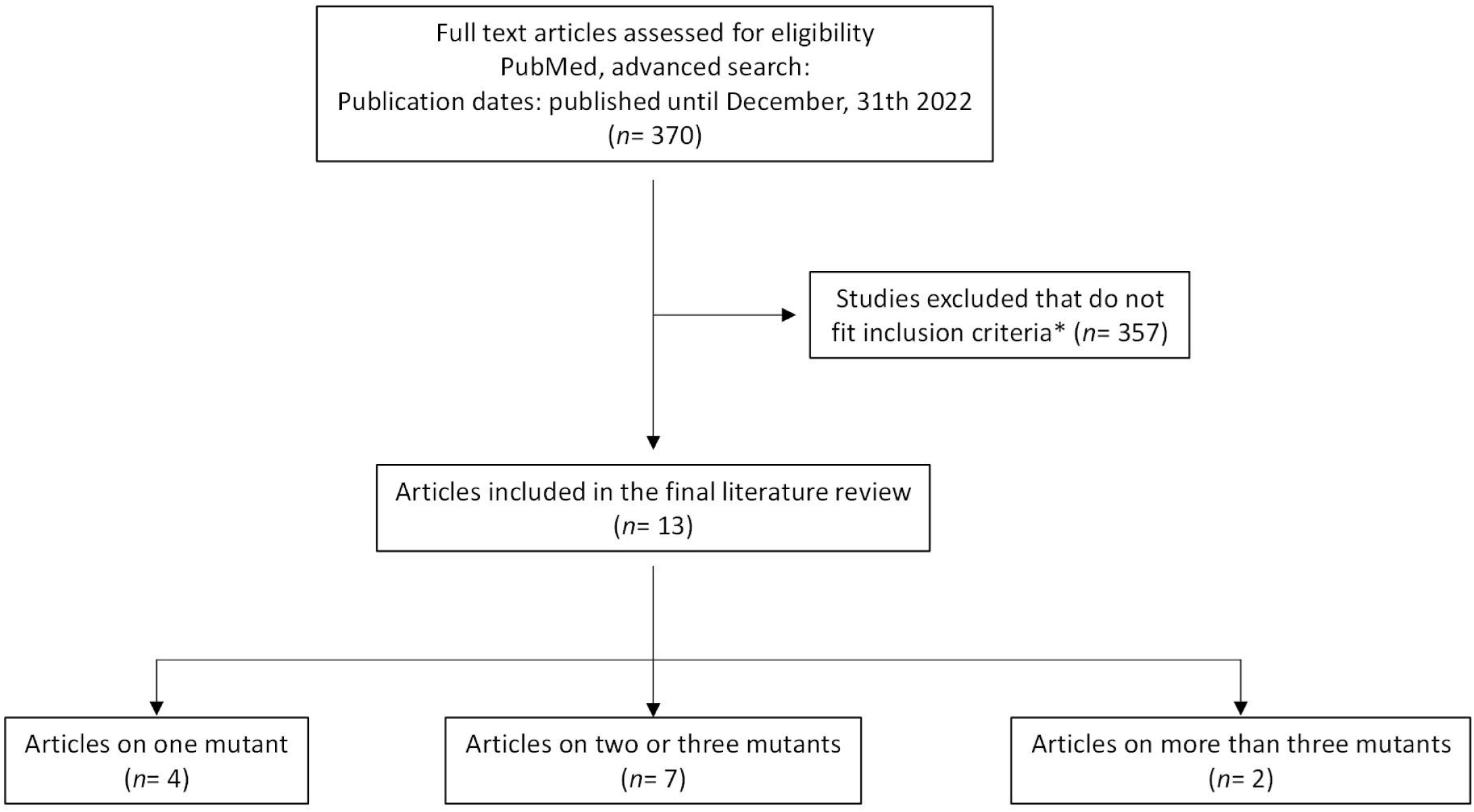
Overview of systematic literature search and study selection process. Systematic literature research was performed in MEDLINE via PubMed including all studies published up to December 31st, 2022. Studies that fit the inclusion criteria were included in the final literature review. *Inclusion criteria encompass published and peer-reviewed studies that analyze the effect of single Omicron-associated spike mutations in the index spike background on neutralization of clinical specimen using neutralization assays. Studies that did not fit the inclusion criteria were excluded.

### Patients and samples

Study participants were recruited in July 2022 in the region of Brunswick in Lower Saxony, Germany. Recruitment of eligible participants (> 18 years) was based on age- and sex-stratified random sampling. Participants were included if they fit into one of the three cohorts: 1) confirmed infection-naïve individuals that received the first booster vaccination, 2) individuals with an Omicron breakthrough infection who received the first booster vaccination, and 3) infection-naïve individuals that received a second booster vaccine based on the index spike. Basic sociodemographic data are self-reported and provided in table 1 and supplementary table 2. Absence of previous infection was self-reported and confirmed using a SARS-CoV-2 nucleoprotein ELISA during sampling. Individuals with an Omicron breakthrough infection were defined as having a positive rapid antigen-test or positive PCR-test after the last vaccination. The infecting virus was defined as the most prevalent strain in Germany at the time of infection. No sequencing was performed. Disease severity was self-reported as mild for all participants with the following predominantly reported symptoms: fever, headache, cough, nasal congestion, fatigue. No included subjects were hospitalized due to COVID-19. Participants were excluded if they had been either previously infected or had more than one breakthrough infection prior to sampling. Peripheral blood was collected from each participant by venipuncture using a serum gel S-Monovette (Sarstedt) and further processed according to manufacturer’s instructions. Serum was then aliquoted and stored at -20°C until further use.

**Table 1.** Cohort characteristics. N/A not applicable. * Vaccines: BNT162b2 (BNT), AZD1222 (VAX), mRN A-1273 (MOD)

| Cohort characteristics | 1<sup>st</sup> Boost, naïve<br>(n = 9) | 1<sup>st</sup> Boost,<br>breakthrough (n = 10) | 2<sup>nd</sup> Boost, naïve<br>(n = 4) |
| --- | --- | --- | --- |
| **Sex, n(%)** |  |  |  |
| Male | 4 (44.4) | 4 (40) | 1 (25) |
| Female | 5 (55.6) | 6 (60) | 3 (75) |
| **Age, median (IQR; range)** | 31<br>(29,53; 25 - 59) | 30<br>(25.8,43.8; 23 - 51) | 27<br>(25.3,33.3; 23 - 49) |
| **Baseline SARS-CoV-2 status, n (%)** |  |  |  |
| Positive | 0 (0) | 10 (100) | 0 (0) |
| Negative | 9 (100) | 0 (0) | 4 (100) |
| Unknown | 0 (0) | 0 (0) | 0 (0) |
| **Vaccine regimen\*, n (%)** |  |  |  |
| BNT+BNT+BNT | 6 (66.7) | 5 (50) | N/A |
| BNT+BNT+MOD | 1 (11.1) | 2 (20) | N/A |
| VAX+VAX+BNT | 1 (11.1) | 0 () | N/A |
| VAX+BNT+BNT | 1 (11.1) | 2 (20) | N/A |
| VAX+BNT+MOD | 0 () | 1 (10) | N/A |
| BNT+BNT+BNT+BNT | N/A | N/A | 3 (75) |
| MOD+MOD+BNT+BNT | N/A | N/A | 1 (25) |
| **Interval in weeks, median (IQR)** |  |  |  |
| Interval dose: 1-2 | 6 (6 - 6.5) | 6 (5.25 - 6) | 5 (4 - 6.3) |
| Interval: dose 2-3 | 28 (26 - 32.5) | 22 (21 - 24) | 26 (24.8 - 31.6) |
| Interval: dose 3-4 | N/A | N/A | 28.3 (20.5 - 34.3) |
| Interval: last dose until serum collection | 32 (31.5 - 35) | 29.75 (28.3 - 32.4) | 3.5 (2.6 - 8.8) |
| Interval: last dose until infection | N/A | 18.5 (13.3 - 23.9) | N/A |
| Interval: infection until serum collection | N/A | 14 (3.1 - 17.6) | N/A |

### Ethics statement

This study is part of the COFONI project Prediction of Escape Mutants (PREMUS) with the title ‘*Verwendung von Proben für die Isolation von anti-SARS-CoV2 Antikörpergenen für die Entwicklung von therapeutischen Antikörpern zur Behandlung von COVID-19*’. Ethical clearing was provided by the approval of the PREMUS project. Before participating in the study, all participants received a letter with information about the study and given suitable time to provide voluntary written informed consent prior to sample collection.

### Cell culture

All cells were cultured at 37°C and 5% CO _2_. VeroE6 (ATCC CRL-1586), and HEK293T (DSMZ ACC-635) cells were cultured in DMEM medium supplemented with 1% Penicillin-Streptomycin, 1% L-glutamine, and 5% fetal bovine serum (FBS).

### Plasmids

The plasmid pCG1_SARS-2-Sdel18 encoding the spike protein of the Wuhan-Hu-1 SARS-CoV-2 (human codon-optimized, 18 amino acid truncation at the C-terminus) has been previously reported (Hoffmann et al., 2020). Plasmids encoding the index SARS-CoV-2 spike protein harboring single Omicron-associated spike mutations were generated via restriction-free cloning. All substitution amino acid changes and single amino acid deletions were integrated using the Q5 Site-Directed Mutagenesis Kit (NEB #E0554S) in the previously mentioned plasmid. All deletions and insertions of multiple amino acids were generated using a mutagenesis PCR and following signature- and ligation-independent cloning (SLIC) reaction. Successful introduction of the respective mutation was verified by sequence analysis using a commercial sequencing service (Microsynth Seqlab). The expression vectors for the SARS-CoV-2 spike of Omicron BA.1 (based on isolate hCoV-19/Botswana/R40B58_BHP_3321001245/2021; GISAID Accession ID: EPI_ISL_6640919), BA.2 (based on isolate hCoV-19/England/PHEC-4G0AFZF7/2021; GISAID Accession ID: EPI_ISL_8738174), BA.2.12.1 (based on isolate hCoV-19/USA/FL-CDC-ASC210848866/2022; GISAID Accession ID: EPI_ISL_12028907), BA.2.75 (based on hCoV-19/England/LSPA-3EAAD0A/2022; GISAID Accession ID: EPI_ISL_13692860), BA.3 (based on isolate hCoV-19/South Africa/NICD-N25677/2021; GISAID Accession ID: EPI_ISL_8801154), BA.4/5 (based on isolate hCoV-19/England/LSPA-3C01A75/2022 [BA.4] and hCoV-19/France/CVL-IPP25260/2022 [BA.5]; GISAID Accession ID: EPI_ISL_11550739 [BA.4] and EPI_ISL_12029894 [BA.5]), BA.4.6 (based on hCoV-19/USA/NE-NPHL22-18369/2022; GISAID Accession ID: EPI_ISL_14429885), BA.5.9 (based on hCoV-19/Denmark/DCGC-549789/2022; GISAID Accession ID: EPI_ISL_13971444), and BF.7 (based on hCoV-19/Denmark/DCGC-550448/2022; GISAID Accession ID: EPI_ISL_13972569) were generated by Gibson assembly as described before (Arora, Kempf, et al., 2022; Arora, Nehlmeier, et al., 2022; Arora, Zhang, Nehlmeier, et al., 2022; Arora, Zhang, Rocha, et al., 2022; Hoffmann et al., 2022). The expression vectors for the SARS-CoV-2 Omicron BQ.1 and BQ.1.1 were generated by introducing spike mutations into the spike of Omicron BA.5 (based on isolate hCoV-19/France/CVL-IPP25260/2022; GISAID Accession ID: EPI_ISL_12029894) via site-directed mutagenesis. The XBB expression plasmid was generated by Gibson assembly based on the expression vector for the spike of Omicron BA.2 (based on isolate hCoV-19/England/PHEC-4G0AFZF7/2021; GISAID Accession ID: EPI_ISL_8738174). Site-directed mutagenesis was utilized to generate the XBB.1 and XBB.1.5 expression plasmids based on the XBB spike expressing plasmid.

### SARS-CoV-2 nucleoprotein ELISA

The Human SARS-CoV-2 Nucleoprotein IgG ELISA Kit (Abbexa) was used according to manufacturer’s instructions. Briefly, the surface of a 96-well plate was coated with the SARS-CoV-2 N-protein and incubated for 45 min at 37°C with 1:100 diluted serum samples heat inactivated at 56°C for 30 min. Plates were washed 3 x with Wash Buffer. Next, the horseradish peroxidase-conjugated anti-human IgG was added and incubated at 37°C for 30 min. Plates were washed 3 x with Wash Buffer and TMB substrate was added to each well and incubated for 20 min at 37°C. After addition of Stop Solution, readout took place using a microplate reader (Tecan Austria GmbH) at 450 nm wavelength. The color intensity value of the negative control was used as a threshold for qualitative identification of SARS-CoV-2 N-protein positive serum samples.

### Production and titration of SARS-CoV-2 pseudo-viruses

Rhabdoviral pseudo types harboring SARS-CoV-2 full or mutant spike proteins were generated as previously described (Becker et al., 2021). Briefly, HEK293T cells were transfected via calcium-phosphate with different SARS-CoV-2 spike protein expression plasmids. After 24h post transfection, cells were transduced using a replication deficient VSV-G (VSV*ΔG) at a multiplicity of infection (MOI) of 3 for 1h at 37°C and 5% CO _2_. After washing with PBS, medium containing an anti-VSV-G antibody (supernatant of L1-hybridoma cells – CRL2700) was added to neutralize residual VSV surface glycoprotein. After incubation for 22h at 37°C and 5% CO _2_, supernatant was pooled, and cellular debris was removed by centrifugation at 2000 g for 5 min at 4°C. Aliquots of the supernatant were stored at -80°C until further use. In addition, pseudo-viruses were titrated on VeroE6 cells to ensure comparable infectivity and to normalize the viral input between assays of all used pseudo-viruses.

### Pseudo-virus neutralization assay

For the assay, all internal controls and serum samples were heat inactivated at 56°C for 30 min. Thawed samples and controls were stored at 4 °C for no longer than 48 h, prior to use. Heat inactivated serum samples and controls were two-fold serially diluted in DMEM [1% Penicillin-Streptomycin, 1% L-Glu, 5% FBS] in a 96-well microtiter plate ranging from 1:10 to 1:5120. An equal volume of SARS-CoV-2 pseudo-viruses was added to the pre-diluted serum samples and incubated for 1h at 37°C and 5% CO _2_. To ensure comparable infectivity of pseudotyped viral particles, a virus input of approximately 300 ffu/ml was intended. The virus-serum sample mixture was then transferred to VeroE6 cells at 100% confluence seeded one day prior to the experiment. After incubation for 24±2 h at 37°C and 5% CO _2_, GFP ^+^ infected cells were counted using an IncuCyte S3 (Sartorius) performing whole-well scans (4x) in phase contrast and green fluorescence settings (300ms exposure). Automated segmentation and counting of fluorescent foci defined as green fluorescent protein GFP ^+^-single cells was performed using the IncuCyte GUI software (versions 2019B Rev1 and 2021B). Raw data was plotted in GraphPad prism version 9.3.1 and pseudo-virus neutralization titer _50_ (PV N_5_T_0_) was calculated using the least squares fit using a variable slope, four-parameter regression analysis. The lower limit of confidence (LLOC) was set at a PVNT_50_ of 10. Non-responders are defined as individuals with a neutralization titer below this threshold. PVNT _50_ values of these non-responders were arbitrarily set to 5 for visualization purpose. All experiments were performed using an internal standard (a pool of six well characterized serum samples of boosted and confirmed infection-naïve individuals), negative controls and virus-input controls to assess the actual virus input of each used pseudo-virus.

### Quantification and statistical analysis

Generation of graphs and statistical calculations were performed in GraphPad Prism version 9.3.1 for Windows (GraphPad Software). The level of statistical significance was defined as: *p < 0.05, ** p < 0.01, *** p < 0.001. The statistical details of each experiment are indicated in the figure legends. All effects on the neutralization titers were analyzed for statistical significance, but only the significant effects were represented in the figures. Differences in neutralization titers between the index pseudo-virus and either Omicron sub-variant pseudo-viruses, or pseudo-viruses harboring additional single mutations in the wild-type spike protein were analyzed using the Brown-Forsythe and Welch ANOVA test without correction for multiple comparisons. Mathematical outliers were defined using the Rout method.

## Results

### Virus input and infection time affect neutralization assay variance

Neutralization assays are widely used to quantify neutralizing antibody titers against SARS-CoV-2. Although these assays are sensitive and reasonably robust within studies, neutralization data often show a high heterogeneity of reported neutralization titers across studies, which limits comparability and translational value. We assessed two technical parameters that are frequently varying across studies, virus input and infection time, to define their role in variability and increase assay robustness. By using varying virus inoculums ranging from approximately 30 to 2000 focus forming units (ffu)/ml in a 96-well format (MOI from 0.0015 to 0.1), we analyzed the impact of virus input on the resulting pseudo-virus neutralization titer _50_ (PVNT_50_) of a polyclonal serum pool. In line with previously published findings (Nie et al., 2020), we observed nominally increasing neutralizing antibody titers, but also a strong increase in variance when using decreasing amount of virus per reaction, with a pronounced inflection point at doses below 325 ffu/ml, corresponding to an MOI of 0.015 (see figure 2a). Applying this to our own study by consequently back titrating all virus inputs for every single measurement when assessing the effect of single spike mutations on neutralization escape, we did not observe a significant effect on variance for a virus input between 100 and 300 ffu/well (MOI between 0.005 and 0.015). A virus inoculum below 100 ffu/well (MOI of 0.005), however, seems to slightly skew neutralization assay results (see supplementary figure 2b).

**Figure 2:**
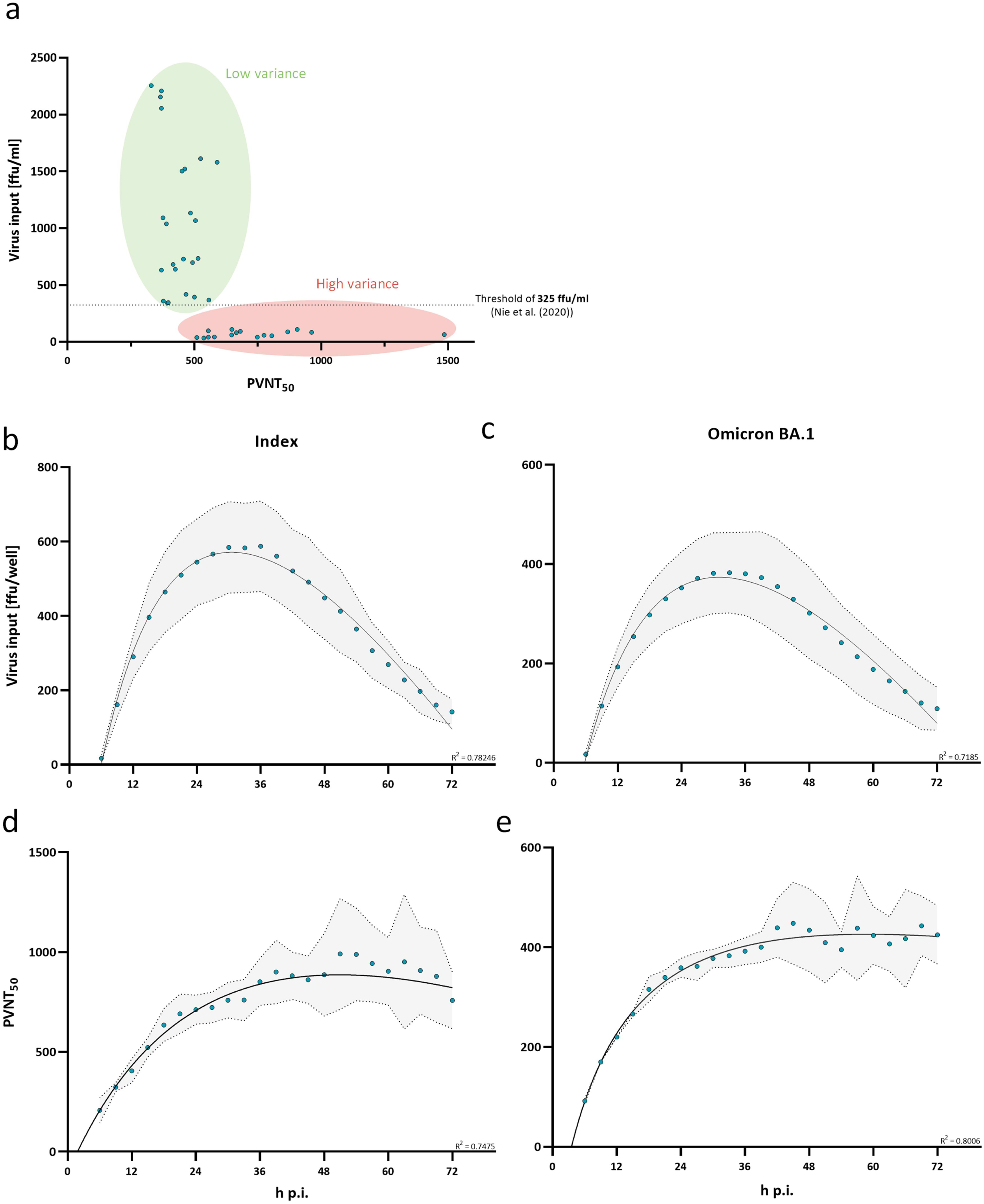
Virus input and infection time affect variance in pseudo-virus neutralization assays. Virus input and infection time were characterized on their ability to affect variance in neutralization assay results. a. Neutralization titers (PVNT50) plotted against the actual virus input. Single points present measurements with virus inputs ranging from 30 to 2500 ffu/ml. A virus input of 325 ffu/ml is indicated by the dashed horizontal line which was determined as a threshold for robust regression fit by Nie et al. (2020). b. and c. Time-dependent differences in the number of detectable infected cells were measured for the index (b) and Omicron BA.1 (c) variant. Single points show the mean measured virus input from three independent experiments with the standard deviation shaded in grey. The relationship between neutralization titer and infection time is described by a second order polynomial (quadratic) regression. R-squared values of the regression are provided in the figure. d. and e. Neutralization titers plotted against infection time (h p.i.) for the index (d) and Omicron BA.1-variant (e). Single points show the mean PVNT50 from three independent experiments with the standard deviation shaded in grey. Relationship between PVNT50 and infection time is described by a second order polynomial (quadratic) regression. R-squared values of the regression are provided in the figure.

We next evaluated the impact of infection time on neutralization titers from pseudo-virus neutralization assays, by performing assays with measurements of infected cells in 3 h intervals for 72 h starting at 6 h post infection. To assess this time-dependent effect, we measured the number of infected cells and the resulting PVNT_50_ over 72 h. We observed that the detectable number of GFP ^+^ infected cells increases during the first 24 h post infection, peaks between 24 and 36 h and then starts to decrease (figure 2b and c). The measured neutralization titer increased during the first 36 h before it started to plateau (see figure 2d and 1e). Importantly, the variance of measured neutralizing antibody titers increased when the virus input started to decrease at around 36 h post infection. Moreover, we used two variants of SARS-CoV-2 spike in the assay, and these observations were independent of the pseudo-virus used (figure 2b and c).

Based on these results, we suggest that similar pseudo-virus based neutralization assays measured between 24 h and 36 h and with a virus input of ≥ 300 ffu/well (MOI of ≥ 0.015) shall be preferentially used to achieve robust and reproducible assay results.

### Diminished neutralization responses towards novel Omicron sub-variants in post-vaccination sera

Since the emergence of the Omicron B.1.1.529 variant in November 2021, new Omicron sub-variants emerged, harboring a plethora of novel spike-mutations (Stanford Coronavirus Antiviral & Resistance Database (CoVDB). We validated the evidence on immune escape of emerging Omicron sub-variants in different clinically relevant immunological settings (figure 3a) in our optimized neutralization assay. Accordingly, we recruited 23 participants that were either three times vaccinated and infection-naïve (*n*= 9) with a median sample collection time of 32 weeks (*IQR*: 31.5 – 35.0) post last vaccination, three times vaccinated and with an any Omicron breakthrough infection (*n*= 10) with a median sample collection time of 29.8 (*IQR*: 28.3 – 32.4) post last vaccination and 14 weeks (*IQR*: 3.1 – 17.6) post infection, or four times vaccinated with a monovalent vaccine and infection-naïve (*n*= 4) with samples taken after 3.5 weeks (*IQR*: 2.6 – 8.8) post last vaccine dose (cohort details see table 1 and supplementary table 1). Neutralization response against the index strain and Omicron sub-variants BA.1, BA.2, BA.2.12.1, BA.2.75, BA.3, BA.4/5, BA.4.6, BA.5.6, BF.7, BQ.1, BQ.1.1, and XBB.1.5 was assessed by pseudo-virus neutralization assays for all cohorts (see figure 3). All participants that received four vaccine doses or experienced breakthrough infection with an Omicron variant, showed detectable neutralizing antibody titers against all analyzed variants. Infection-naïve participants that received only one booster vaccination (three vaccine doses) showed detectable titers against the index, Omicron BA.1, BA.2, BA.2.75, BA.3, and BF.7 strains. Neutralizing titers against Omicron BA.2.12.1, BA.4/5, BA.4.6, and BA.5.9 were detected in 88.9% of samples. Neutralization response to novel variants like Omicron BQ.1, BQ.1.1, and XBB.1.5 was even lower with 55.6%, 66.7%, and 33.3% responders, respectively. The strongest immune escape was observed in the most recently circulating variant XBB.1.5 with a 98.7-, 68.2-, and 134.6-fold change in neutralizing antibody titers compared to the index strain in the boosted and infection-naïve, boosted and breakthrough infected, and the infection naïve and twice boosted cohort, respectively. Omicron breakthrough infection in participants who previously received three vaccine doses increased the neutralization capacity towards novel variants. On the other hand, infection-naïve individuals whose immune response is only based on vaccine-elicited antibodies against the index spike show poor neutralization of these antigenically distinct variants, especially in the context of waning immunity (see figure 3b). A second booster dose encoding the index antigen boosted overall neutralization responses but failed to induce robust neutralization responses against currently circulating Omicron sub-variants.

**Figure 3:**
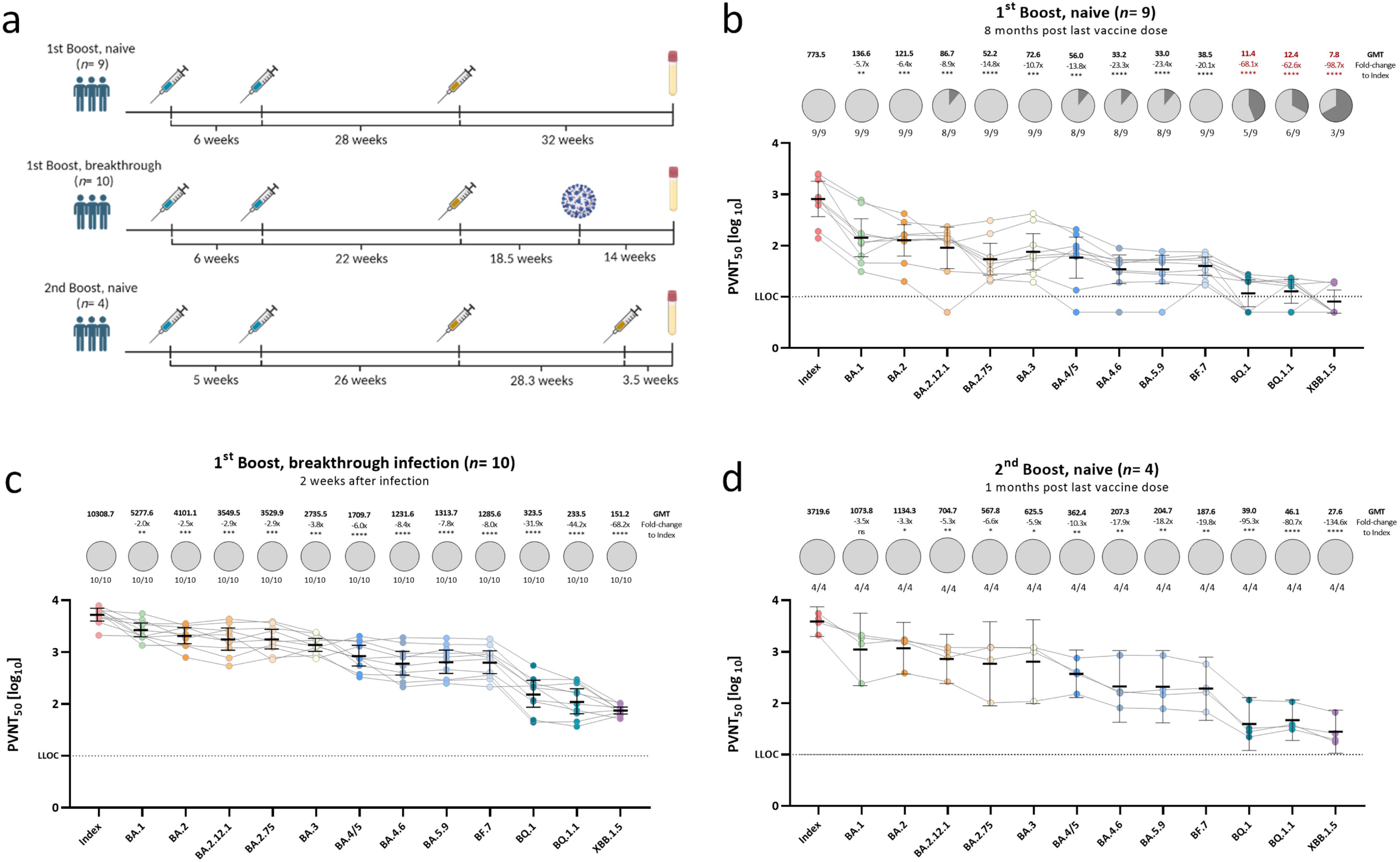
Diminished neutralization responses towards novel Omicron sub-variants in individuals with varying immunological backgrounds. Vaccination- and/ or hybrid immunity-induced neutralizing antibody titers against SARS-CoV-2 index, or Omicron BA.1, BA.2, BA.2.12.1, BA.3, BA.4/5, BA.2.75, BA.4.6, BA.5.9, BF.7, BQ.1, BQ.1.1, and XBB.1.5 pseudo-viruses. a. Schematic presentation of vaccination or infection events of each assessed cohort. All vaccine doses were with monovalent formulations of the index strain. b. Individuals that received either homologous or heterologous AZD1222 and/ or BNT162b2 two-dose primary regimen with an index-based mRNA booster vaccination without previous infection (n= 9), c. Individuals that received either homologous or heterologous AZD1222 and/ or BNT162b2 two-dose primary regimen with an index-based mRNA booster vaccination and an Omicron breakthrough infection after the last vaccination (n= 10), d. Individuals that received homologous mRNA-1273 or BNT162b2 primary regimen and two BNT162b2 index-based booster doses without previous infection (n= 4). PVNT50 data is expressed for each serum sample using a logarithmic scale. Bold horizontal lines and whiskers are mean PVNT50 ± standard deviation. Geometric mean titers (GMTs) are shown above the individual measurements. Fold-changes in neutralization capacity between index and the respective Omicron sub-variant together with the statistical significance is shown below the GMTs. GMTs and fold-changes that contain >20% of non-responders are highlighted in red. Dashed line indicates the lower limit of confidence (LLOC) at a PVNT50 of 10. Statistical analysis was performed by the Brown-Forsythe and Welch ANOVA test without correction for multiple comparisons. Statistical significance was defined by a value of *<0.05; ** <0.01; *** <0.001.

### Systematic assessment of single Omicron-associated spike mutations on neutralization escape

To understand the molecular basis of the strong immune escape seen in clinically relevant Omicron sub-variants, we systematically evaluated the impact of every single Omicron-associated spike mutation on serum neutralization. First, we performed a systematic literature research on available studies assessing neutralization of the SARS-CoV-2 index virus harboring single Omicron-associated spike mutations using clinical specimen. For this, the spike mutations of the Omicron sub-variants BA.1, BA.2, BA.2.12.1, BA.3, and BA.4/5 were assessed (figure 4a). Until the 14^th^ of April 2023, 8 out of 46 spike mutations (except for the mutation D614G) were not at all evaluated on their specific effect on immune escape using clinical specimen (see figure 4b). One study comprehensively assessed spike mutations of Omicron BA.1 and BA.2, but not Omicron BA.2.12.1, BA.3, or BA.4/5. In this study, Pastorio et al. (2022) measured neutralizing antibody titers against pseudo-viruses harboring BA.1 and BA.2 mutations in the index spike protein against six individual serum samples. Most other mutations that were described by more than one study are either conserved throughout the Omicron variants (G339D, S371F, K417N, T478K, N501Y, P681H, N764K, Q954H, N969K) or were present in the earliest Omicron sub-variant BA.1 (A701V, L981F). A systematic biological characterization of neutralization escape of all spike mutations of Omicron sub-variants up to BA.5, however, was to the best of our knowledge not performed so far. To close this gap, we produced VSV pseudo-viruses with the spike protein of the index strain harboring each singular Omicron spike mutation and measured their impact on neutralization escape using a polyclonal post-vaccination, infection-naive serum pool. A serum pool enables a focused analysis of the impact of spike mutations on a relatively broad spectrum of post-vaccination specimen, without additional variance caused by individual responses. All pseudo-virus stocks were titrated to ensure comparable infectivity and to normalize the viral input between assays of all used pseudo-viruses. Since pseudo-virus titers were determined by their ability to infect VeroE6 cells following standardized protocols, resulting titers can be interpreted as a proxy for viral infectivity. For most mutants, we did not observe a significant effect on virus titers as compared to the index virus. However, some mutants resulted in consistently lower titers as compared to the index and other full-spike pseudo-viruses of Omicron sub-variants (figure 5a). We show that the mutations Δ69-70, S371F, S371L, S375F, T376A, Q496S, and N501Y on their own affect pseudo-virus infection of VeroE6 cells. It was reported previously that the three mutations in the serine residues S371F, S371L, and S375F that are part of a core subdomain clustering at a hairpin loop, as well as the adjacent T376A mutation significantly affect spike protein expression and processing in pseudo-virus particles possibly due to impairment of the main-chain conformational change (Lan et al., 2022; Pastorio et al., 2022). Because virus stocks of these variants could not be produced with sufficient titers to infect target cells at comparable MOI, they were not used for the assessment of neutralization and where therefore excluded from the main experiment. When assessing neutralization escape of the remaining single-mutation constructs, we identified several single-mutations with a significant effect on antibody neutralization of our serum pool (figure 5b). As expected, mutations in the RBD had the strongest effect on spike function, and among five tested mutations, G339D and D405N resulted in a 1.5-fold (*SD* = 0.2) increased neutralization escape compared to the index strain, while the mutation K417N enhanced susceptibility to neutralization (1.8-fold (*SD* = 0.2)). Two more mutations N679K and P681H present in the subdomain 1 and 2 were also shown to result in significant neutralization resistance with a 1.6-fold (*SD* = 0.4; 0.3) increase in neutralization escape, whereas the mutation H655Y in the SD1/2 domain resulted in a significant 1.6-fold (*SD* = 0.2) increase of neutralization sensitivity compared to the index strain. No mutations with a significant effect on neutralization were identified in the NTD or RBM. As expected, the Omicron full-spike variants showed the highest fold-reduction of neutralization titers in our serum pool relative to the index strain with up to 16.1-fold (*SD* = 6.2) increased titers for Omicron BA.4/5.

**Figure 4:**
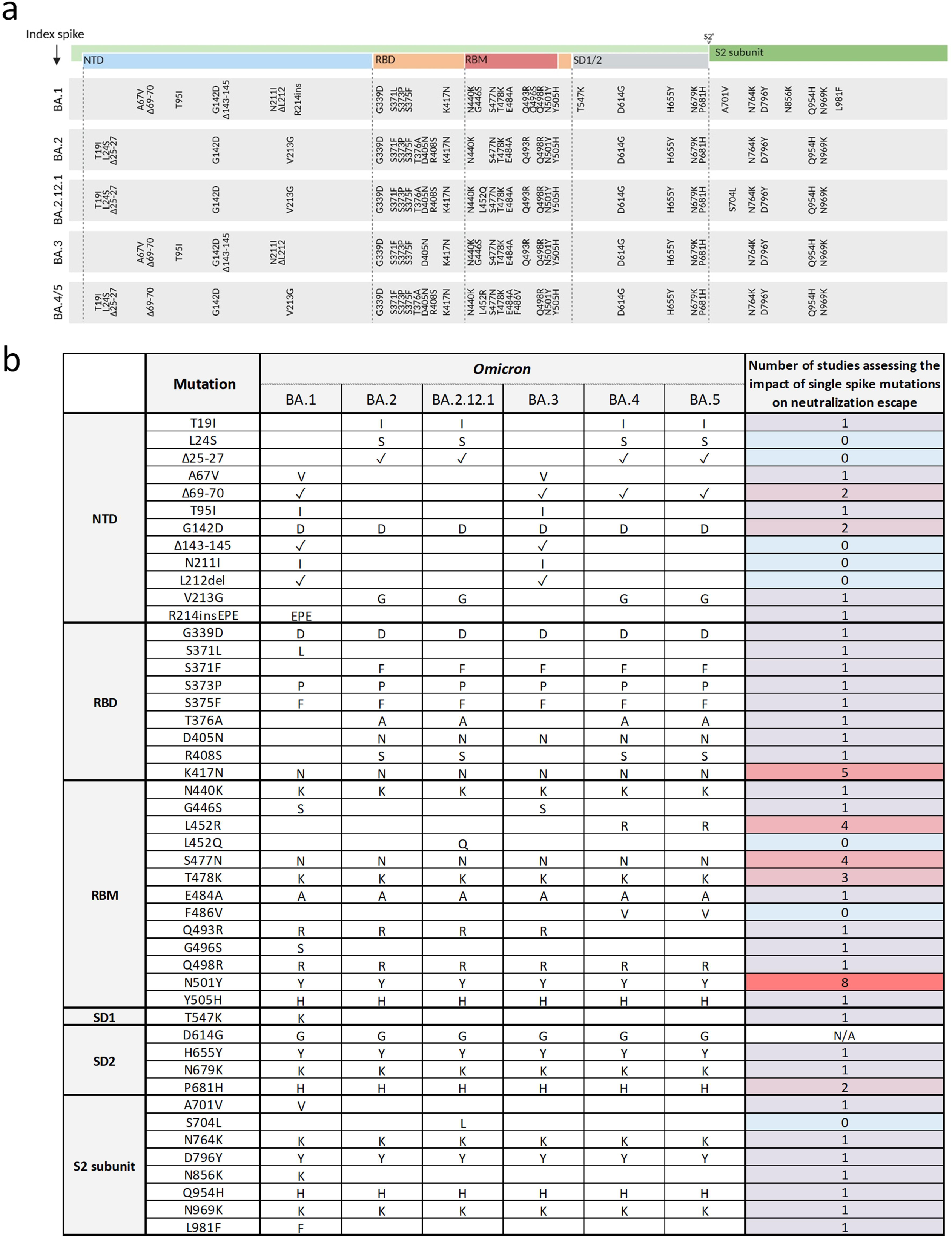
Overview of Omicron spike mutations and corresponding evidence on their effect on neutralization by clinical specimen. a. Schematic presentation of the SARS-CoV-2 spike domains and amino acid changes indicated for Omicron variants BA.1, BA.2, BA.2.12.1, BA.3, and BA.4/5 compared to the index spike. N-terminal domain (NTD, blue); receptor-binding domain (RBD, orange); receptor-binding motif (RBM, red); subdomains 1 and 2 (SD1/2, grey); S2 cleavage site (S2’); S2 subunit (dark green). Mutations were obtained from the Stanford Coronavirus Antiviral & Resistance Database (CoVDB). b. Overview of the number of studies analyzing single Omicron SARS-CoV-2 spike mutations on their effect on neutralization by clinical specimen. Mutations are organized according to their position in the spike protein domains: NTD, RBD, RBM, SD1/2, and S2 subunit. Presence of the mutation in the respective Omicron sub-variant is either indicated by the substituted or inserted amino acids, or the presence of a deletion as indicated with a tick. The evidence on the observed biological effects of each mutation is displayed and color-coded based on the number of studies assessing the mutation. b. Fold-reduction of the mean PVNT50s from VOC (dark petrol) and single mutant (petrol) pseudo-viruses relative to the index strain were measured in a pool of six serum samples from boosted infection-naive individuals. Raw neutralization data are provided in the supplements. Mutants with significant fold-reductions are shown in red. Mutants with potentially biased significant fold-reductions are shown in light petrol. No data was obtained for mutants with too low virus titers because of technical limitations (indicated as n.d.). Respective spike domains of mutants are indicated below the x-axis. Bars and whiskers present the mean fold-reduction ± standard deviation. Standard deviation of the index fold-reductions are shown as a light grey horizontal bar. Single dots represent single measured fold-reductions. Significant fold-reductions, as well as significance levels are provided on the top of the graph. Statistical analysis was performed using the Brown-Forsythe and Welch ANOVA test without correction for multiple comparisons. Statistical significance was defined by a value of * < 0.05; ** < 0.01; *** <0.001. n.s. is not significant; n.a. is not available. c. Heat map showing the fold-reduction in neutralization titers of single Omicron-associated spike mutations with their occurrence in the respective Omicron sub-variant. Numbers are fold-reductions in neutralization titers relative to the index strain ± standard deviation.

**Figure 5:**
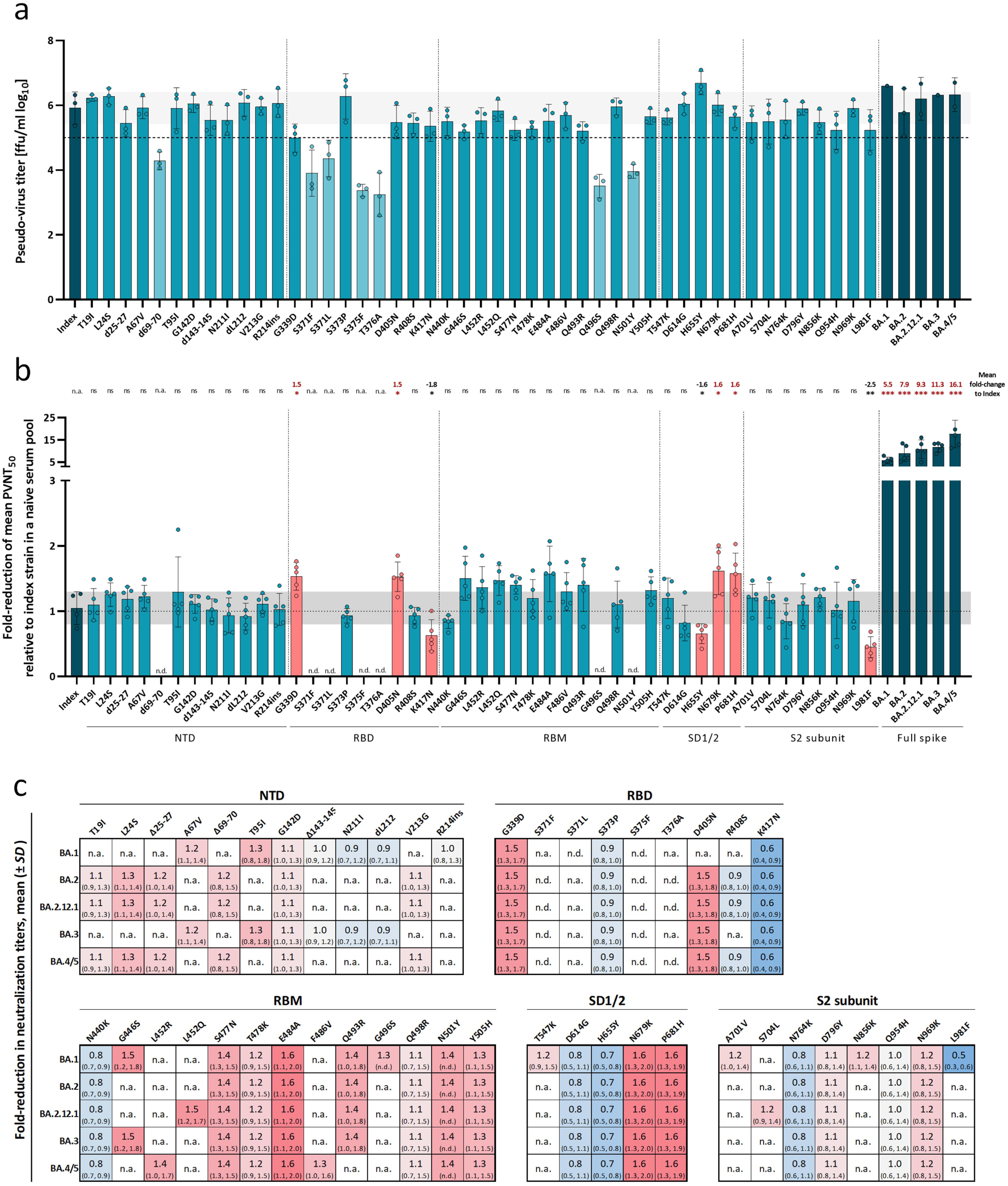
Impact of Omicron-associated spike mutations on infectivity and serum neutralization. Effect of single Omicron-associated spike mutations was characterized by their ability to affect infection of VeroE6 cells and neutralization capacity from vaccination-elicited neutralizing antibodies compared to the index strain. a. Overview of pseudo-virus titers determined by VeroE6 cell infection of all produced full spike VOC and single mutant VSV-pseudo-viruses. Mean titers from pseudo-viruses with single Omicron-associated mutations in the index S background (petrol) and pseudo-viruses of VOCs (dark petrol) are shown. Dashed horizontal line indicates a titer of 105 ffu/ml; pseudo-viruses with titers below this threshold are shown in light petrol. Columns and whiskers are mean pseudo-virus titers ± standard deviation. Single dots represent independent technical replicates. Standard deviation of the index strain is shown as a light grey horizontal bar. Respective SARS-CoV-2 spike domains are indicated below the x-axis.

Notably, most point mutations that result in a significant fold-change were conserved throughout the analyzed Omicron sub-variants (see figure 5c). Interestingly, the L981F mutation with a significant fold-change of 0.5 (*SD* = 0.2) relative to the index strain, which indicates an increased susceptibility towards neutralization was only present in Omicron BA.1, but not in the following sub-variants. Other mutations strongly reducing susceptibility towards neutralization like G339D, N679K, and P681H are present in all analyzed clinically relevant Omicron sub-variants indicating an important advantage for the virus.

### Defective viral particles potentially impact pseudo-virus neutralization assay results

Some mutations resulted in strongly reduced pseudo-virus infectivity and could only be grown to low titers. Pseudo-virus neutralization assays with these mutants (Δ69-70, S371L and S375F) show dramatically increased neutralization escape with a fold-reduction in neutralizing antibody titers of up to 3.7- and 7.8-fold relative to the index strain respectively for S371L and S375F (see supplementary figure 2a). Surprisingly and suspiciously, the strength of this effect was comparable to the escape observed for Omicron BA.1 and BA.2. Since it is unrealistic to assume that these mutations alone could confer this magnitude of resistance, we assumed that these results may have been biased by technical artifacts.

Structurally important mutations like S371L and S375F could lead to improper folding and significantly diminished functionality of spike. When pseudo-viruses carrying these mutations are produced and titrated, titrated stocks would likely contain significant amounts of dysfunctional particles not being identified during titration, because these particles fail to infect and to induce expression of marker genes. Nevertheless, these mutated spike proteins might still harbor epitopes that can be bound by neutralizing antibodies during neutralization assays. Consequently, these defective particles could act as a sponge for antibodies and deter the neutralization of the remaining infectious particles causing an artificial decrease in observed neutralization effects.

To explore this hypothesis, we performed neutralization assays using an index pseudo-virus with different proportions of UV-inactivated index virus to mimic defective viral particles not causing GFP expression. Virus stocks with varying percentages of inactivated virus were titrated and diluted to a consistent input of 300 ffu/well (MOI of 0.015) of functional virus (see supplementary figure 1a and b). When measuring neutralizing antibody titers with these virus preparations, we observed significant reductions of neutralizing antibody titers that correlated with increasing amounts of defective viral particles (see supplementary figure 1c). 90% inactivated virus resulted in a 1.9-fold drop in neutralizing antibody titer relative to 0% of defective viral particles. This result indicated that high proportions of defective viral particles may skew neutralization assay results towards a lower neutralization titer. This relationship can possibly be described by an asymptotic curve which indicates that a low quantity of defective viral particles has no significant effect on the resulting neutralization titer, whereas a higher ratio of defective vs. infective viral particles results in a higher influence on the neutralization titer. Hence, the strong escape from neutralization that was observed for pseudo-viruses with strikingly low titers after production were likely due to defective viral particles influencing the PVNT _50_ and not to immune escape properties of the mutation per se.

## Discussion

We reported recently that neutralizing antibody data are highly heterogeneous across studies even when cohort characteristics were similar, which indicates that technical considerations can alter the outcome of neutralization assays across studies, limiting their translational value (Jacobsen et al., 2022). To increase assay robustness and data comparability, we assessed the infection-dose and -time as two important technical factors possibly affecting assay variance. We observed that inconsistent infection times result in inconsistent neutralization titers, and hence, heterogeneous data. This conclusion especially applies to the readout of pseudo-virus based assay systems that rely on marker gene expression. Furthermore, we show that the infectious dose also affects neutralization results. Higher virus doses led to nominally lower neutralization titers, because more virus can infect the cells without being neutralized by antibody-containing serum. On the other side, lower doses of input virus do not only increase the nominal neutralization titers observed in assays, but also the variance in observed titers, resulting in less precise measurements. This effect is most pronounced for virus inputs below 300 ffu/well (MOI < 0.015), in line with a previous report (Nie et al., 2020). To increase robustness of neutralization assay results, we strongly advise the usage of a consistent viral inoculum ≥ 300 ffu/well (MOI ≥ 0.015) and measurements at a uniform time post infection between 24h and 36h, which in our hands resulted in lower variance in neutralizing antibody titers.

Applying these optimized assay conditions, we analyzed the neutralization capacity of sera from vaccinated and hybrid-immune individuals against clinically relevant and newly emerging Omicron sub-variants up to the currently circulating variant XBB.1.5 mostly confirming existing evidence. By trying to understand the molecular basis of this strong immune escape, we identified distinct mutations that were able to significantly impact neutralization escape. In total, we examined 47 mutant pseudo-viruses harboring amino acid changes in the index spike background from Omicron spikes of BA.1, BA.2, BA.2.12.1, BA.3, and BA.4/5. We observed fewer mutations significantly affecting neutralization than observed for monoclonal antibody neutralization possibly due to the usage of a polyclonal serum pool of three times vaccinated individuals. We found the RBD mutations G339D and D405N, as well as SD1/2 mutations N679K and P681H to significantly increase neutralization escape relative to the index strain by 1.5- to 1.6-fold respectively. These mutations have, to our knowledge, not been described in their potential to affect the neutralization response towards polyclonal sera. Previous reports, however, show that G339D can escape broad sarbecovirus neutralizing antibodies which is further increased by synergistic mutations (Cao et al., 2022). These interactions of multiple mutations were also shown to increase levels of infectivity as seen for polybasic furin cleavage site (FCS) mutations P681H with N501Y, as well as for the mutation A701V in combination with N501Y relative to the index strain (Kuzmina et al., 2022). The positioning of P681H adjacent to the FCS could also indicate a role in spike protein processing and hence, increased infectivity (Jaimes et al., 2020). In addition, we identified the three spike mutations K417N, H655Y, and L981F that significantly increase susceptibility to neutralization by polyclonal post-vaccination sera by 1.6- to 2.5-fold. It was previously shown that K417N reduces ACE2 binding affinity (Laffeber et al., 2021; Mannar et al., 2021), and increases susceptibility to plasma neutralization as confirmed in this study (a Wang et al., 2021; b Wang et al., 2021). In combination with other mutations like N501Y and E484K, however, K417N contributes to a strong increase in viral infectivity and binding capacity (Li et al., 2021).

We also identified spike mutations that strongly affect the functionality of the spike protein. The mutations Δ69-70, S371F, S371L, S375F, T376A, N501Y, and G496S showed reduced infectivity of VeroE6 cells. In addition, the two mutants S371L and S375F led to abnormally high fold-reductions of neutralizing antibody titers relative to the index strain that were even comparable to the fold-reductions of Omicron BA.1 and BA.2 relative to the index. By crystal structure analysis, it was found that the mutations S371L/F, S373P, and S375F are part of a core subdomain clustering at a hairpin loop which results in a main-chain conformational change (Lan et al., 2022). Consequently, we assume that the presence of singular mutations in this region could impair proper spike protein folding and consequently increase the chances of interfering defective viral particle formation. This observation could be extended to the adjacent T376A mutation as it was previously seen in Pastorio et al. (2022) that mutations at these serine residues led to severely diminished infection of CaCo2 cells. In addition, they report a highly reduced incorporation of S375F and T376A harboring spikes, and reduced processing of spikes containing the mutations S371F and S371L. Accordingly, diminished cell-entry could be explained by impaired binding to the ACE2-receptor by either single mutation-induced conformational changes, or overall reduced spike protein stability. We have shown in a correlative experiment that high fractions of defective viral particles can significantly affect downstream applications. The relationship between the percentage of defective viral particles and neutralization titers might be possibly described as an asymptotic curve. Consequently, the neutralization titers of pseudo-viruses with very low virus titers after production need to be treated with caution, because they may contain a high fraction of defective particles, whereas the neutralization titers of pseudo-viruses with comparable infectious titers are likely to be minimally affected by this concern. Of course, there could be additional causes of the low infectivity of VeroE6 cells that we observed in this study for the mutations Δ69-70 and G496S that are less likely attributed to structural reasons and need further examination.

Taken together, we identified critical factors in pseudo-virus neutralization assays that potentially affect assay readouts and hinder intra-experimental comparability of data. In addition, we generated a comprehensive library of pseudo-viruses encoding all single spike mutations of clinically relevant Omicron sub-variants until BA.4/5 and applied this library to characterize the ability of each mutation to alter viral susceptibility to neutralization by polyclonal post-vaccination serum. This pseudo-virus library may also be used for studies of single mutations in the context of monoclonal antibody development or in other applications. However, further studies are necessary to understand the functionality of each spike mutation in the context of Omicrons immune escape, and their possible implications on the future development of vaccines and therapeutics.

## Limitations

We did not cover the impact of combinations of spike mutations present in Omicron on the neutralization by polyclonal post-vaccination sera in this study. Singular spike mutations could behave differently in terms of infectivity and neutralization escape than in the context of a full-functional spike. In addition, data resulting from pseudo-viruses, rather than replication-competent recombinant SARS-CoV-2 viruses, may only serve as a proxy for the neutralization escape and viral infectivity. Furthermore, the relatively small cohort size, especially for the four times vaccinated individuals, used in the analyses of immune escape of newly emerging Omicron sub-variants limits the strength of the conclusions that may be inferred from it. We also did not assess the potency of bivalent booster vaccines in the neutralization of novel Omicron sub-variants. Lastly, we performed the neutralization assays using virus with an intended MOI of 0.015, however, the de facto input slightly varied between experiments. Of our assays 25.4% were performed using an MOI < 0.015 and 5.8% were below 0.005 (see supplementary figure 2). While we identified a higher variance in data of experiments with an MOI < 0.005, this was not the case for data of experiments with MOIs > 0.005. Hence, some outlier data points could be attributed to increased variance by lower viral inoculate, but there is no significant consensus explaining the outliers.

## Data Availability

All data produced in the present study are available upon reasonable request to the authors

## Acknowledgements

We thank all study participants for their commitment and enthusiasm to contribute to this study. We thank Daniela Lenz, Ayse Barut, and Inge Hollatz-Rangosch for excellent technical assistance during this study. We thank the Peter and Traudl Engelhorn foundation for providing a post-doctoral fellowship to HJ. We thank Gert Zimmer (Institute of Virology and Immunology, Mittelhäusern, Switzerland) for providing the VSV pseudo-virus system.

## Authors contributions

M.K. performed the experiments with assistance of D.C.C. and under supervision of H.J. H.J. L.C.S and M.K conceived the study. M.K. and H.J. performed data analysis and interpretation. M.K. generated figures and tables. J.vdH supervised SLIC-adapted cloning experiments. M.H. and S.P. provided expression plasmids. H.G. conceived the ethical permit. M.K. and H.J. wrote the manuscript. LCS acquired funding. All authors critically reviewed and approved the final manuscript.

## Lead contact

Further information and requests for resources and reagents should be directed to and will be fulfilled by the lead contact, Henning Jacobsen.

