## Supplementary material for "Systematical assessment of the impact of single spike mutations of SARS-CoV-2 Omicron sub-variants on the neutralization capacity of post-vaccination sera"

### Supplementary information

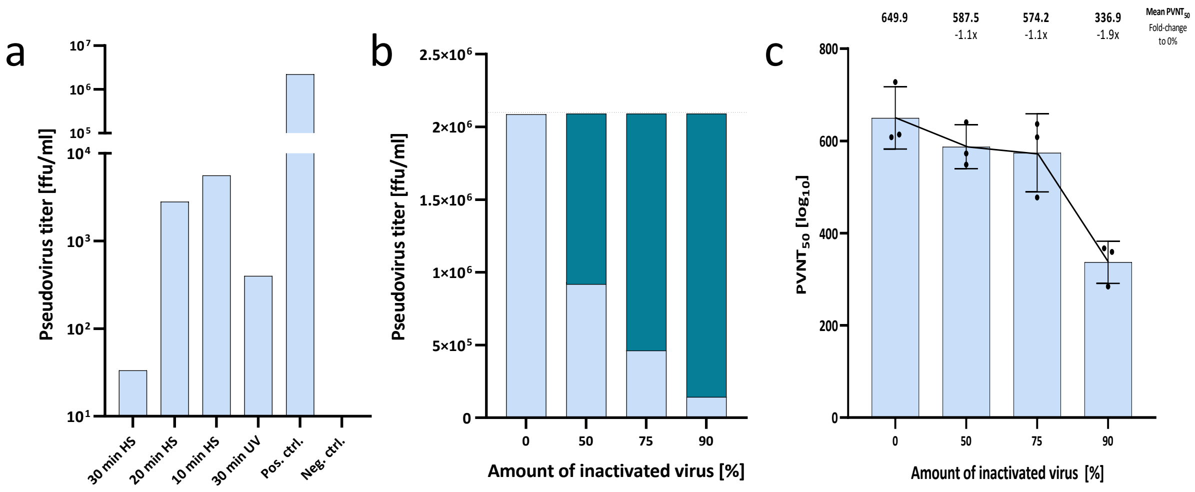


**Supplementary figure 1**

Pseudo-virus titers measured after different inactivation protocols and using varying amounts of inactivated virus. **a**. shows the pseudo-virus titers in ffu/ml after heat (HS)- or UV-inactivation. Bars present the total pseudo-virus titer determined by the number of infected VeroE6 cells. **b**. shows the pseudo-virus titer of pseudo-virus stocks containing 0%, 50%, 75%, or 90% of UV-inactivated index pseudo-virus (light blue). The pseudo-virus titer that was supposedly reduced by UV-inactivation is shown as petrol columns for each virus stock up to the titer of the index virus stock with 0% UV-inactivated virus (indicated as dashed horizontal line). **c**. $PVNT_{50}$s against index pseudo-virus containing equal infective viral particles with 0%, 50%, 75%, or 90% of UV-inactivated index pseudo-virus. Mean $PVNT_{50}$s are plotted and shown on top of the graph. Fold-change relative to the virus containing no UV-inactivated index virus is shown below the $PVNT_{50}$s. Bars and whiskers present the mean $PVNT_{50}$ ± standard deviation. Single dots indicate average neutralization titers from three independent technical replicates.

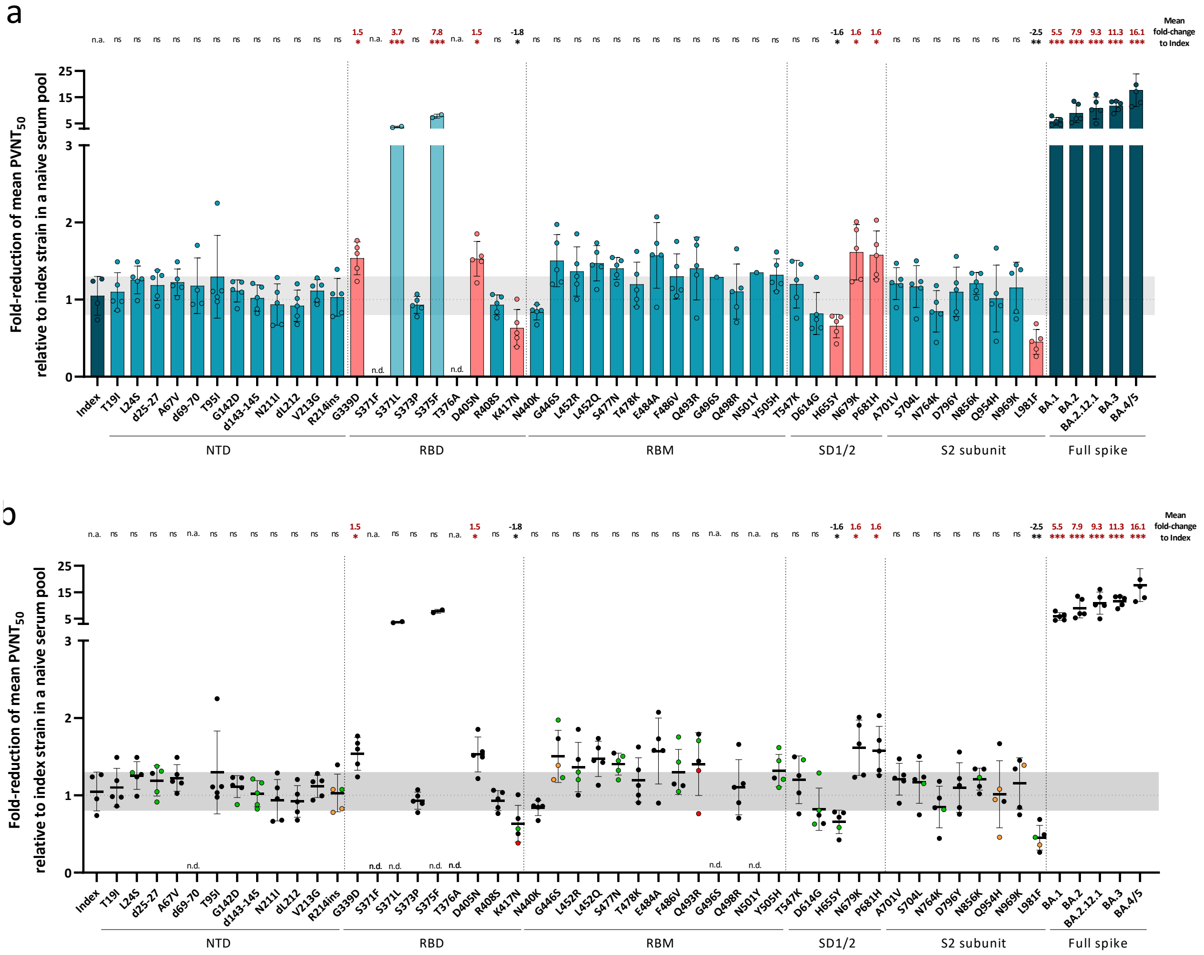


**Supplementary figure 2**

**a**. Fold-reduction of the mean PVNT$_{50}$s from VOC (dark petrol) and single mutant (petrol) pseudo-viruses relative to the index strain were measured in a pool of six serum samples from boosted infection-naive individuals. Raw neutralization data are provided in the supplements. Mutants with significant fold-reductions are shown in red. Mutants with potentially biased significant fold-reductions are shown in light petrol. No data was obtained for mutants with too low virus titers because of technical limitations (indicated as n.d.). Respective spike domains of mutants are indicated below the x-axis. Bars and whiskers present the mean fold-reduction ± standard deviation. Standard deviation of the index fold-reductions are shown as a light grey horizontal bar. Single dots represent single measured fold-reductions. Significant fold-reductions, as well as significance levels are provided on the top of the graph. Statistical analysis was performed using the Brown-Forsythe and Welch ANOVA test without correction for multiple comparisons. Statistical significance was defined by a value of * < 0.05; ** < 0.01; *** <0.001. n.s. is not significant; n.a. is not available. **b**. shows identical data to a. but with single PVNT$_{50}$s color coded according to the quantity of virus input used in each experiment. Virus input > 300 ffu/well (black), 200 – 300 ffu/well (green), 100 – 200 ffu/well (orange), < 100 ffu/well (red).

**Supplementary table 1** Detailed participant characteristics. N/A not applicable; BNT162b2 (BNT), AZD1222 (VAX), mRNA-1273 (MOD)

| | Sex | Age group | Baseline SARS-CoV-2 status | Vaccine type | Interval: dose 1-2 | Interval: dose 2-3 | Interval: dose 3-4 | Interval: last dose until serum collection | Interval: last dose until infection | Interval: infection until serum collection | Severity (WHO grade) | Prevalent variant | Date of infection |
|---|---|---|---|---|---|---|---|---|---|---|---|---|---|
| 1st Boost, naive | m | 30-40 | negative | BNT+BNT+BNT | 3 weeks | 42 weeks | N/A | 29 weeks | N/A | N/A | N/A | N/A | N/A |
| | f | 30-40 | negative | VAX+BNT+BNT | 10 weeks | 28.5 weeks | N/A | 32 weeks | N/A | N/A | N/A | N/A | N/A |
| | f | 50-60 | negative | VAX+VAX+BNT | 10 weeks | 26 weeks | N/A | 32 weeks | N/A | N/A | N/A | N/A | N/A |
| | f | 20-30 | negative | BNT+BNT+BNT | 6 weeks | 32.5 weeks | N/A | 29 weeks | N/A | N/A | N/A | N/A | N/A |
| | m | 50-60 | negative | BNT+BNT+MOD | 6 weeks | 22 weeks | N/A | 35 weeks | N/A | N/A | N/A | N/A | N/A |
| | f | 20-30 | negative | BNT+BNT+BNT | 4 weeks | 45.7 weeks | N/A | 39 weeks | N/A | N/A | N/A | N/A | N/A |
| | f | 50-60 | negative | BNT+BNT+BNT | 6 weeks | 25.5 weeks | N/A | 31.5 weeks | N/A | N/A | N/A | N/A | N/A |
| | m | 20-30 | negative | BNT+BNT+BNT | 6 weeks | 28 weeks | N/A | 31.5 weeks | N/A | N/A | N/A | N/A | N/A |
| | m | 50-60 | negative | BNT+BNT+BNT | 6.5 weeks | 26 weeks | N/A | 36.5 weeks | N/A | N/A | N/A | N/A | N/A |
| 1st Boost, breakthrough | f | 30-40 | positive, no PCR | BNT+BNT+MOD | 3 weeks | 21 weeks | N/A | 28 weeks | 13 weeks | 16 weeks | Mild | BA.2 | March 2022 |
| | f | 20-30 | positive (confirmed) | BNT+BNT+BNT | 6 weeks | 21 weeks | N/A | 29.5 weeks | 1.5 weeks | 28 weeks | Mild | BA.1/ BA.2 | January 2022 |
| | f | 20-30 | positive, no PCR | BNT+BNT+BNT | 6 weeks | 21 weeks | N/A | 30 weeks | 26 weeks | 3.5 weeks | Mild | BA.5 | June 2022 |
| | f | 20-30 | positive (confirmed) | VAX+BNT+BNT | 12 weeks | 24 weeks | N/A | 29 weeks | 6 weeks | 24 weeks | Mild | BA.1/ BA.2 | January 2022 |
| | m | 30-40 | positive, no PCR | BNT+BNT+BNT | 6 weeks | 28 weeks | N/A | 26.5 weeks | 23.5 weeks | 3 weeks | Mild | BA.5 | July 2022 |
| | m | 30-40 | positive (confirmed) | VAX+BNT+BNT | 11.5 weeks | 29 weeks | N/A | 28 weeks | 24 weeks | 3 weeks | Mild | BA.5 | July 2022 |
| | f | 50-60 | positive (confirmed) | BNT+BNT+MOD | 6 weeks | 20 weeks | N/A | 30.5 weeks | 14 weeks | 16.5 weeks | Mild | BA.2 | March 2022 |
| | m | 50-60 | positive (confirmed) | VAX+BNT+MOD | 6 weeks | 20 weeks | N/A | 33 weeks | 28 weeks | 3 weeks | Mild | BA.5 | July 2022 |
| | m | 20-30 | positive, no PCR | BNT+BNT+BNT | 4 weeks | 24 weeks | N/A | 37 weeks | 21 weeks | 12 weeks | Mild | BA.2 | March 2022 |
| | f | 30-40 | positive, no PCR | BNT+BNT+BNT | 5 weeks | 23 weeks | N/A | 40 weeks | 16 weeks | 18 weeks | Mild | BA.2 | March 2022 |
| 2nd Boost, naive | m | 20-30 | negative | MOD+MOD+BNT+BNT | 6 weeks | 24 weeks | 10 weeks | 23 weeks | N/A | N/A | N/A | N/A | N/A |
| | f | 20-30 | negative | BNT+BNT+BNT+BNT | 7 weeks | 25 weeks | 24 weeks | 4 weeks | N/A | N/A | N/A | N/A | N/A |
| | f | 50-60 | negative | BNT+BNT+BNT+BNT | 4 weeks | 27 weeks | 32.5 weeks | 3 weeks | N/A | N/A | N/A | N/A | N/A |
| | f | 20-30 | negative | BNT+BNT+BNT+BNT | 4 weeks | 45.7 weeks | 39.5 weeks | 1.5 weeks | N/A | N/A | N/A | N/A | N/A |

**Supplementary table 2** Study characteristics of all studies from the systematic literature review.

| Study First Author | Journal | Date | Analyzed mutations | Specimen |
|---|---|---|---|---|
| Pastorio[1] | Cell Host & Microbe | 14.09.2022 | G339D, S373P, S375F, K417N, N440K, S477N, T478K, E484A, Q493R, Q498R, N501Y, Y505H, H655Y, N679K, P681H, N764K, D796Y, Q954H, N969K, A67V, d69-70, T95I, d142-144, R214EPE, S371L, G446S, G496S, T547K, N856K, L981F, T19I, G142D, V213G, S371F, T376A, D405N, R408S | Serum |
| Wang[2] | Nature | 10.02.2021 | K417N, N501Y | Plasma |
| Cao[3] | Nature | 21.05.2021 | K417N, N501Y | Plasma/ serum |
| Wang[4] | CellPress Immunity | 13.07.2021 | K417N, N501Y | Plasma |
| Neerukonda[5] | Viruses | 11.12.2021 | T478K, K417N, L452R | Serum |
| Alenquer[6] | PLOS Pathogens | 05.08.2021 | N501Y, S477N, L452R, | Serum |
| Ferreira[7] | Journal of Infectious Diseases | 09.09.2021 | L452R | Serum |
| Zhang[8] | Emerging Microbes & Infections | 07.04.2022 | L452R, T478K, G142D | Serum |
| Wang[9] | Emerging Microbes & Infections | 21.12.2021 | L452Q | Serum |
| Wu[10] | Frontiers | 17.06.2021 | S477N | Serum |
| Kuzmina[11] | Viruses | 13.04.2022 | P681H, A701V, N501Y, L452R, S477N | Serum |
| Tada[12] | American Society for Microbiology | 01.06.2021 | d69-70, N501Y | Plasma |
| Xie[13] | Nature medicine | 08.02.2021 | N501Y | Serum |
